# Evaluation and comparison of nine growth- and development-based measures of pubertal timing

**DOI:** 10.1101/2023.06.12.23290796

**Authors:** Ahmed Elhakeem, Monika Frysz, Ana G Soares, Joshua A Bell, Tim J Cole, Jon Heron, Laura D Howe, Sylvain Sebert, Kate Tilling, Nicholas J Timpson, Deborah A Lawlor

**Author notes:** Correspondence: MRC Integrative Epidemiology Unit at the University of Bristol, Oakfield House, Oakfield Grove, Bristol BS8 2BN, UK.

## Abstract

Puberty timing is fundamentally part of life-course health. Yet, little is known about the value of different measures of pubertal timing, particularly in males. We used a population-based cohort to examine nine measures of pubertal age (n=8,500), identifying development of pubic hair in males (12.6y) and breasts in females (11.5y) as early indicators of puberty, and voice breaking (14.2y) and menarche (12.7y) as late indicators. All measures showed evidence of positive phenotypic intercorrelations (e.g., r=0.49: male genitalia and pubic hair ages), and positive genetic intercorrelations. A genetic risk score (GRS) for age at menarche associated positively with all other measures (e.g., difference in female peak height velocity age per SD higher GRS: 0.24y, 95%CI: 0.21 to 0.26), as did GRS for voice breaking age (e.g., difference in male axillary hair age: 0.11y, 0.07 to 0.15). We illustrate the value of different pubertal age measures and their use in life-course research.

---

Puberty is a milestone in human development that involves rapid transformations in anatomy, physiology, and behaviour. Its central feature is neuroendocrine transformation of processes regulating reproductive physiology via a reactivation of the hypothalamic-pituitary-gonadal (HPG) axis, leading to onset of adult reproductive capacity^1, 2^. Reactivation of the HPG axis produces numerous observable downstream consequences including production of gonadal steroids, a pubertal growth spurt, development of secondary sexual characteristics, onset of menstruation in females, and the appearance of facial hair and voice change in males^2, 3^. The sequence in which observable changes appear is thought to mirror elevation of steroid levels, with all changes occurring earlier in females than males^2^.

There is substantial variation in the age of puberty between children^4, 5^, which is attributable to genetic and non-genetic factors, such as nutrition^6–9^. Understanding the determinants of the variation in puberty timing is important given its relation to reproductive capability and social and health implications, including risk of some cancers^8–16^. Most research relating to puberty timing has relied on reported age at menarche (a notable singular event, with no clear male equivalent) and therefore has been undertaken in females only. Other measures of pubertal age are available, including those that can be used in both sexes (e.g., height-based growth measures)^17–19^. However, to the best of our knowledge, no study has systematically compared anthropometric and developmental measures of pubertal age. A detailed analysis of different measures of pubertal age in both sexes can help reveal their sequence, interrelationships, and value for future research and data collection strategies.

The aim of this study was to evaluate and compare multiple measures of puberty timing. We used a UK birth cohort – the Avon Longitudinal Study of Parents and Children (ALSPAC)^20–,22^ – where offspring have been prospectively assessed since birth with extensive biomedical data collections that included repeated assessments of height, weight, and bone in research clinics, and repeated assessments of pubertal development using questionnaires. Importantly, assessments began at age 7 years, i.e., before onset of puberty in most children. We derived nine anthropometrical and developmental measures of pubertal age in 8,500 females and males, describe timing and chronological sequence of pubertal growth and development, the phenotypic and genetic correlations between the measures of pubertal age, and how each pubertal age measure relates to genetic risk scores (GRSs) for puberty timing and adiposity, and phenotypic measurements of child body composition.

## RESULTS

### Characteristics of the study participants

Measures of pubertal age were estimated for up to 4,267 females and 4,251 males who had completed at least one of up to nine repeated research clinic assessments (from mean age 7.6 to 17.8 years) where bone mineral content (BMC), height, and weight were recorded; or at least one of up to nine repeated puberty questionnaires (mean ages 8.2 to 17.0 years) where menarche, Tanner stages, and axillary hair and voice breaking status were reported (**Fig. 1**, **Supplementary Table 1**, **Supplementary Table 2**). When compared with those included in estimation of pubertal age, those excluded due to missing data on all clinic and questionnaires assessments had younger maternal age at birth, lower maternal education, higher prevalence of maternal pregnancy smoking, mothers who were likely to have had previous pregnancies resulting in live birth, similar maternal pre-pregnancy BMI, and somewhat higher childhood energy intake (**Supplementary Table 3**).

**Fig. 1.**
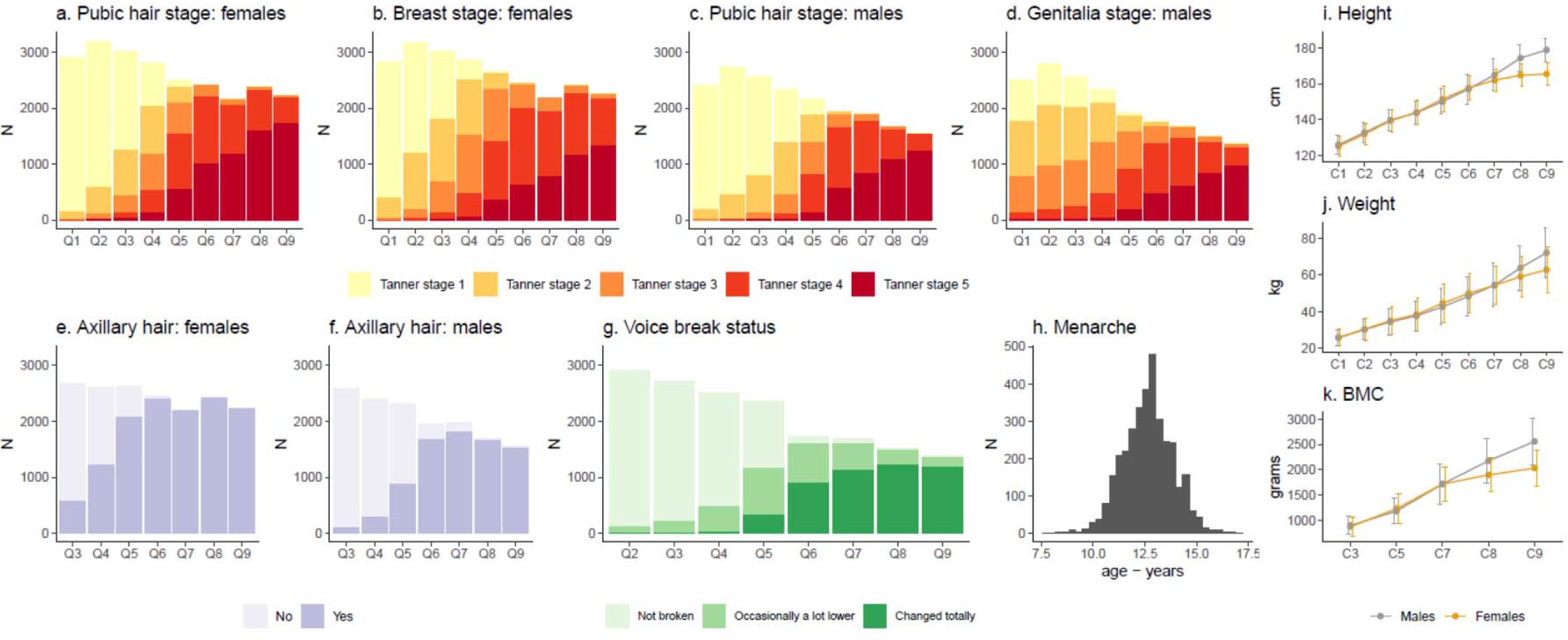
Longitudinal pubertal growth and development data used to derive nine measures of pubertal age. Figure shows number of study participants in each Tanner (pubic hair, breast, genitalia) stage, axillary hair, and voice breaking group at each puberty questionnaire (Q), the distribution of age at menarche, and the mean (± SD) height, weight, and bone mineral content (BMC) at each research clinic (C). Data were collected from mean age 7.6 to 17.8 years; mean age at completing each puberty questionnaire and research clinic assessment is shown in Supplementary Table 2.

### Timing, sequence, and duration of pubertal growth and development

A total of nine measures of pubertal timing (estimated as age in months and presented in years to aid interpretation) were derived. These consisted of two measures in females only (age at menarche and age in Tanner breast stage 3), two in males only (age at voice breaking and age in Tanner genitalia stage 3), and five in both females and males (age at peak BMC velocity, age at peak height velocity, age at peak weight velocity, age in Tanner pubic hair stage 3, and age at axillary hair). Except for age at menarche, which was calculated as the first reported age, all pubertal age measures were derived by applying mixed effects models to each set of repeated puberty assessments (**Fig 1**) and identifying the age corresponding to the peak of the velocity curve^23, 24^.

Mean age of puberty in females varied across measures from 11.5 years for age in Tanner breast stage 3) to 12.7 years for age at menarche (average of 1.2 years from earliest to latest measure), and in males from 12.6 years for age in age in Tanner pubic hair stage 3 to 14.2 years for age at voice break (average of 1.6 years from earliest to latest measure) (**Fig. 2**). The largest time gap between mean ages of consecutive measures was 0.3 years for females (from Tanner pubic hair stage 3 to axillary hair, and from peak BMC velocity to menarche) and 0.7 years for males (from Tanner genitalia stage 3 to peak weight velocity). There was considerable variability between individuals in the age of puberty, e.g., standard deviations around the mean ages ranged in females from 0.8 years (peak height velocity) to 1.2 years (menarche), and in males from 0.7 years (peak BMC velocity) to 1.2 years (Tanner genitalia stage 3, peak weight velocity). Mean age of puberty was younger in females than males for all five measures common to both sexes, e.g., 11.8 years versus 13.5 years for age at peak height velocity (**Fig. 2**).

**Fig. 2.**
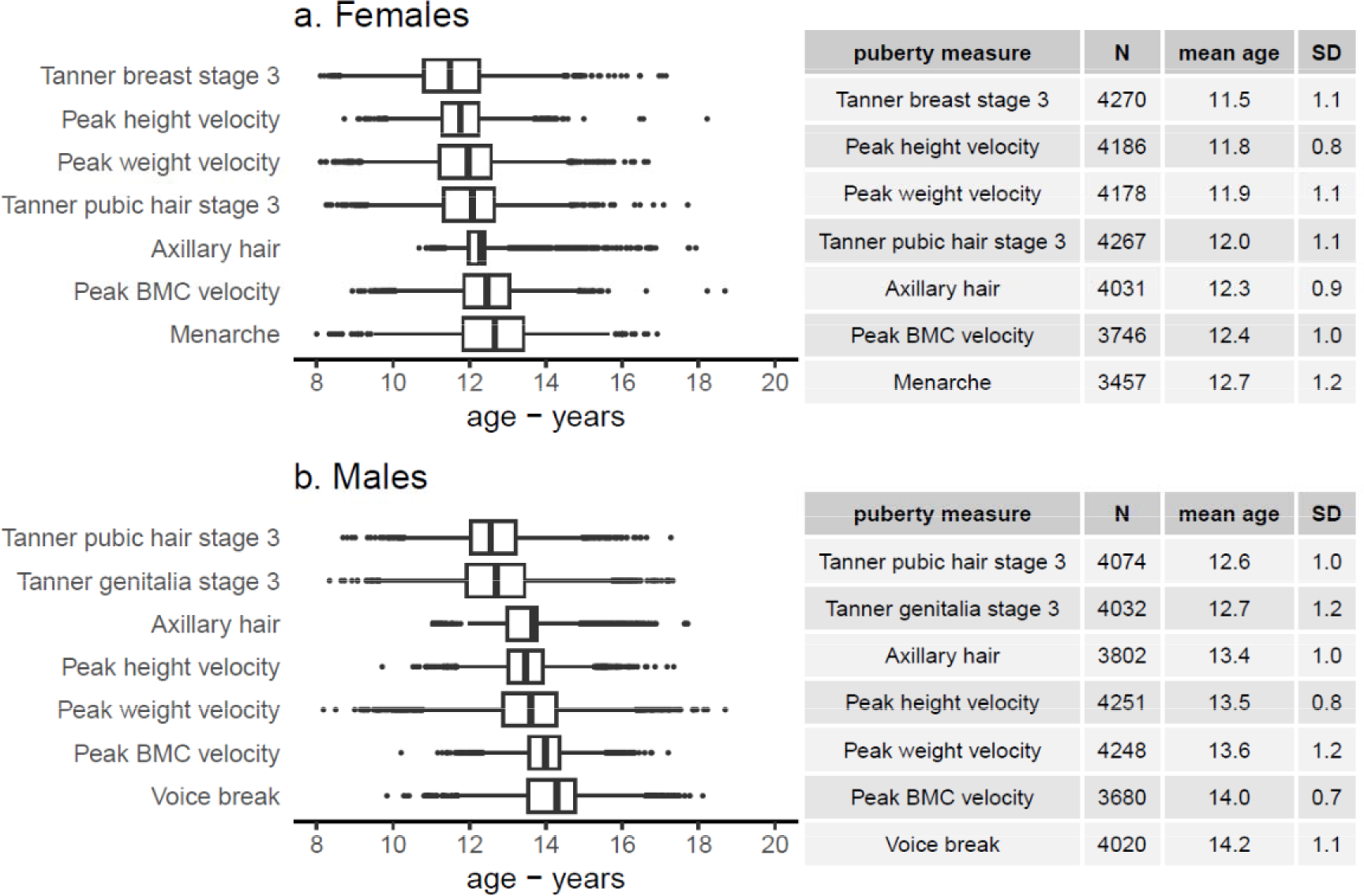
**Timing of pubertal growth and development**. Figure shows summary statistics for the nine pubertal age measures in females and males. The left panels are box plots showing the median, 25^th^ and 75^th^ centiles, and minimum and maximum values of age, plus any outliers for each puberty measure. The right panels show the sample size along with the mean and SD of age (years) for each puberty measure. The measures are arranged by chronological sequence from youngest to oldest.

### Phenotypic correlations between pubertal age measures

Pair-wise phenotypic (Pearson) correlation analyses identified positive, generally moderate strength, correlations between all puberty age measures, with mainly stronger correlations in females (**Fig 3**) than males (**Fig 4**). In females, correlations ranged from 0.28 (between age at axillary hair and age at peak weight velocity) to 0.76 (age at menarche and age at peak height velocity). In males, correlations were from 0.17 (between age in Tanner genitalia stage 3 and age at peak weight velocity) to 0.77 (age at peak height velocity and peak BMC velocity).

**Fig. 3.**
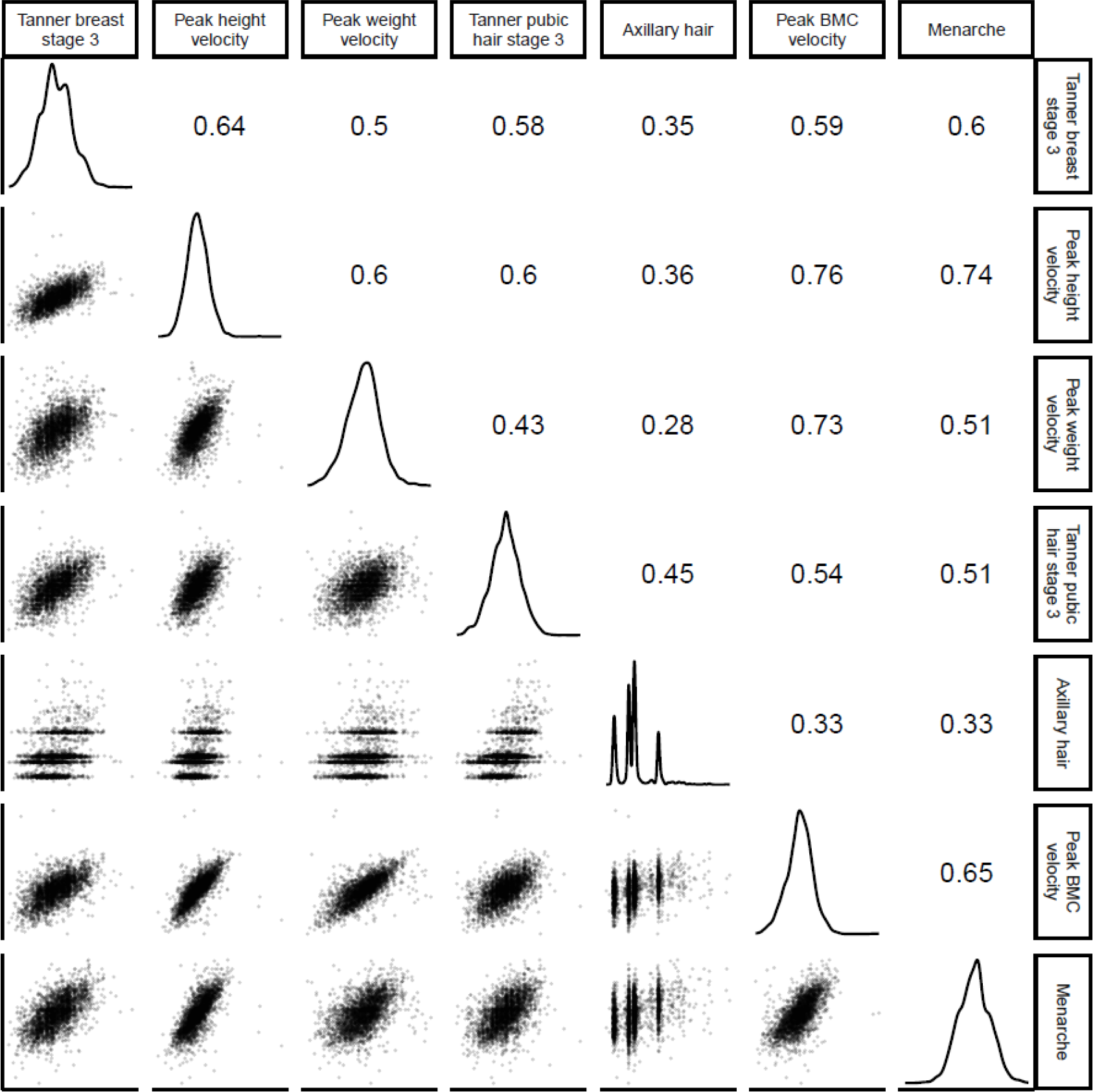
Phenotypic correlations between measures of pubertal age in females. Figure shows pairwise scatterplots (lower triangle), and Pearson correlations (upper triangle) between pubertal age measures, and density plots (diagonal) for each measure.

**Fig. 4.**
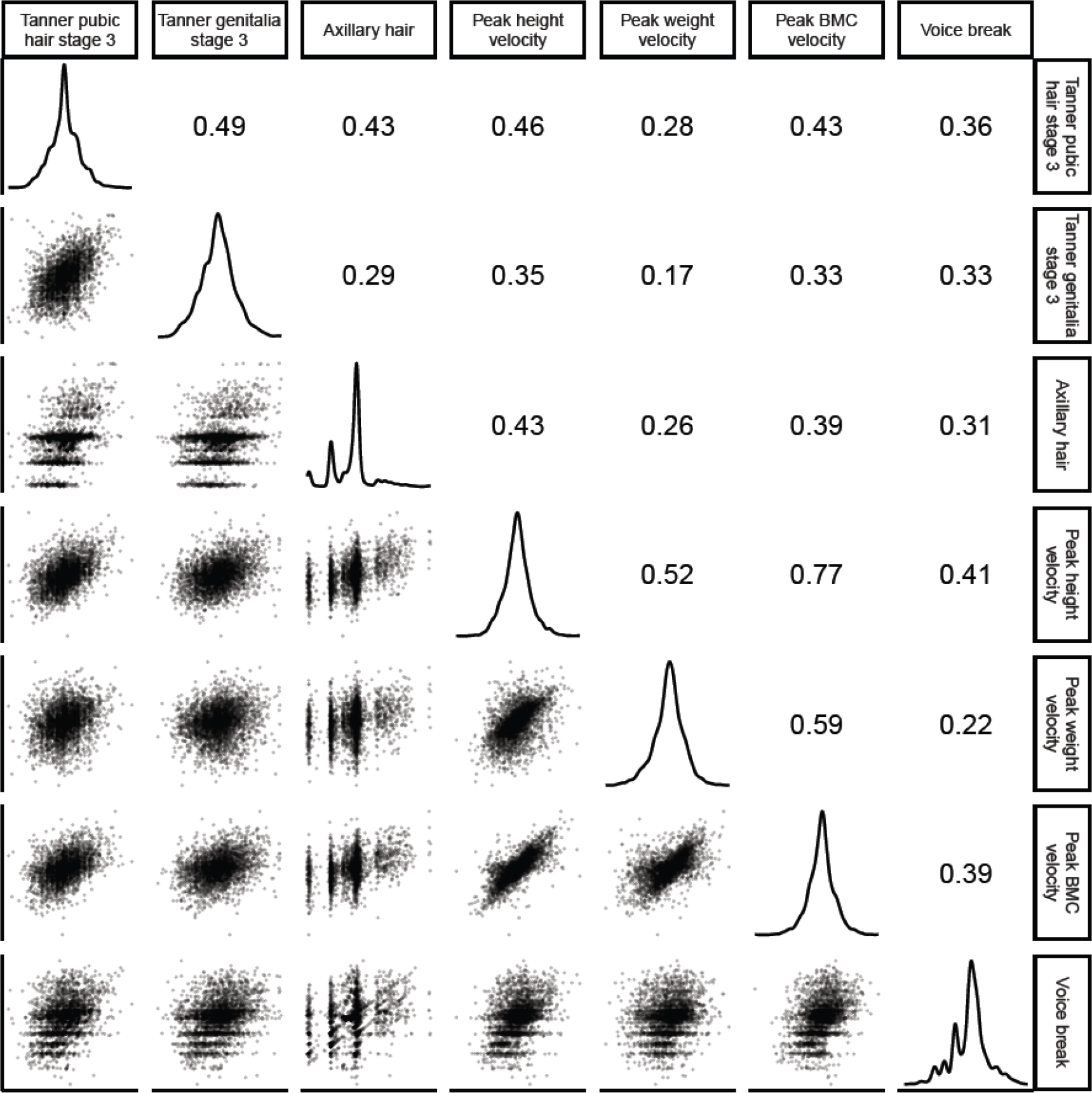
Phenotypic correlations between measures of pubertal age in males. Figure shows pairwise scatterplots (lower triangle), and Pearson correlations (upper triangle) between pubertal age measures, and density plots (diagonal) for each measure.

### Genetic correlations between pubertal age measures

We used linkage disequilibrium score regression (LDSR) to investigate the shared genetic correlations between all pubertal age measures both within and between sexes using genome-wide association study (GWAS) summary statistics. Full summary data were available from a published GWAS on age at menarche^8^ (n=252,000) and were used here. Summary data for all other measures (including for voice breaking since full summary data were not available from the published GWAS on age at voice breaking^9^) were generated using sex specific GWAS in ALSPAC (including follow-up GWAS meta-analyses for the five measures that are available for females and males), ALSPAC GWAS sample sizes ranged from 3,109 (age at peak BMC velocity in males) to 6,782 (age at peak height velocity in females and males combined).

LDSR revealed mostly moderate to high genetic correlations between measures of pubertal age (**Supplementary Table 4**). This included genetic correlations between measures within each sex (for example, genetic correlation between age in Tanner pubic hair stage 3 and age at axillary hair in females was 0.87, P=0.002), and between sex: both within measures (for example, genetic correlation between females and males for age at peak height velocity was 0.64, P=0.05) and across different measures (for example, genetic correlation between age at menarche in females and age at peak BMC velocity in males was 0.78, P=0.007).

### Associations of genetic risk scores (GRSs) for puberty timing and adiposity with pubertal age measures

To evaluate the usefulness of our derived pubertal age measures, we used univariable linear regression models to regress each derived pubertal age measure on four standardised GRSs that were constructed from published genome-wide significant SNPs for puberty timing^8, 9^ and adiposity^25, 26^. Higher GRSs which were associated with older female and male puberty were both associated with older age of all derived pubertal age measures, and higher GRSs which were associated with higher adulthood and childhood BMI were both associated with younger age of all derived pubertal age measures, except for age in Tanner genitalia stage 3 (**Fig. 5**). The associations of puberty timing GRSs with pubertal age measures were generally stronger for the female puberty timing GRS in females and were similar in magnitude for both scores in males. Associations of adulthood and childhood BMI GRSs were similar in magnitude for both scores in both females and males (**Fig. 5**).

**Fig. 5.**
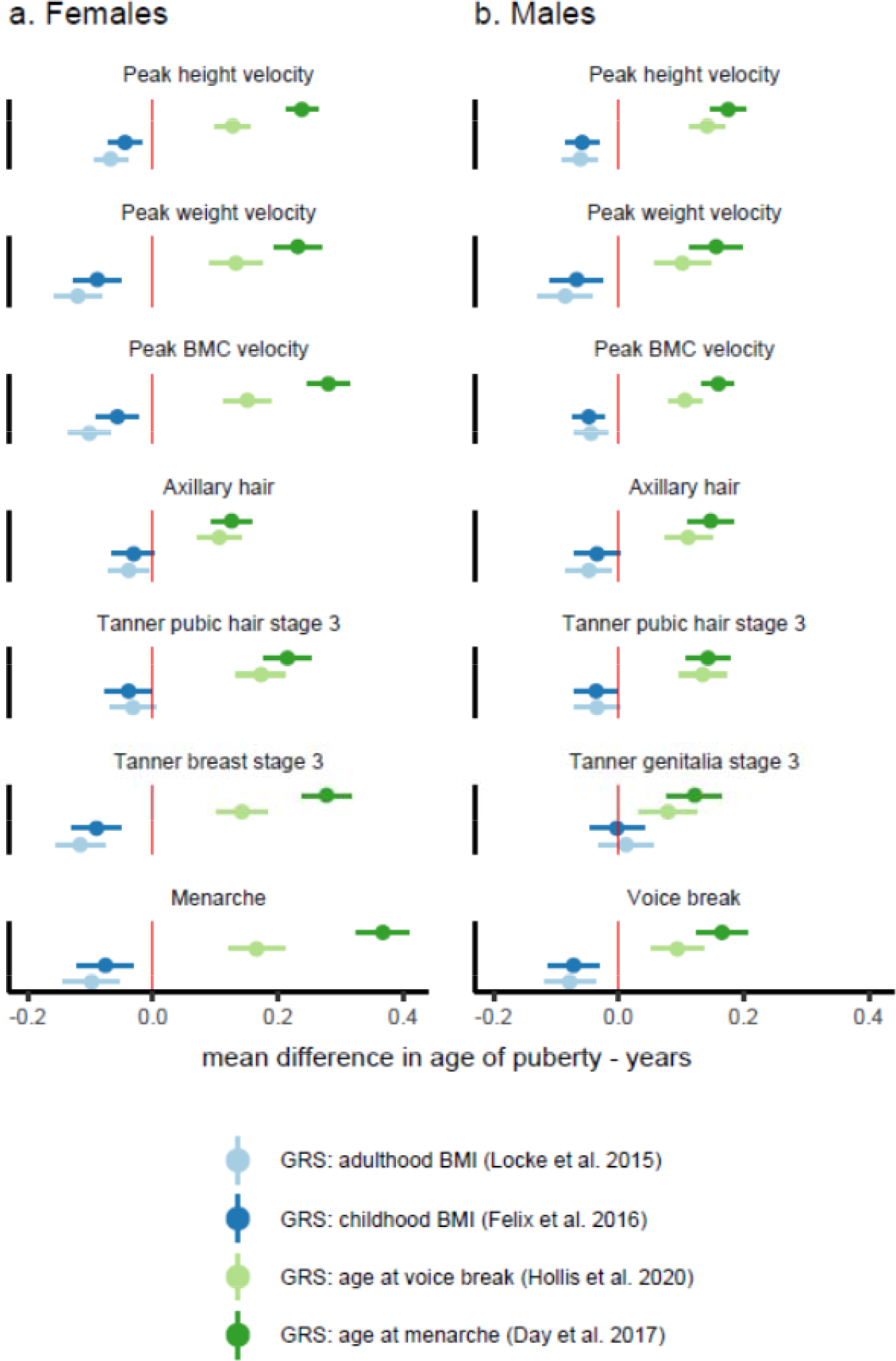
Association between genetic risk scores (GRS) and measures of pubertal age. Figure shows mean difference in age of each puberty measure per standard deviation increase in GRS (for age at menarche, age at voice break, adulthood body mass index (BMI), childhood BMI). Estimates were obtained from separate linear regression models for each pubertal age measure and each GRS. Horizontal bars represent 95% confidence intervals.

### Association of pre-pubertal body composition with puberty age measures

We investigated effects of childhood body composition (Dual-energy X-ray Absorptiometry (DXA) assessments of fat mass and lean mass indices at mean age 10 years), on our derived measures of pubertal age using multivariable linear regression models adjusted for measured confounders (maternal age at birth, maternal pregnancy BMI, maternal pregnancy smoking, maternal education, parity, childhood dietary intake), and exact age at measurement of fat / lean mass. DXA measures recorded after age of puberty were removed, leaving up to 2,491 females and 2,500 males for analysis. Higher pre-pubertal fat mass and lean mass indices were associated with younger age of all puberty measures in females and males. The only exception was for age in Tanner genitalia stage 3 in males, where a higher fat mass index associated with older age (**Fig 6**). Associations of fat mass and lean mass with measures of pubertal age were mostly similar in magnitude in females and were stronger for fat mass in males. Associations with age at peak weight velocity were noticeably stronger for fat mass than lean mass in both females and males (**Fig 6**).

**Fig. 6.**
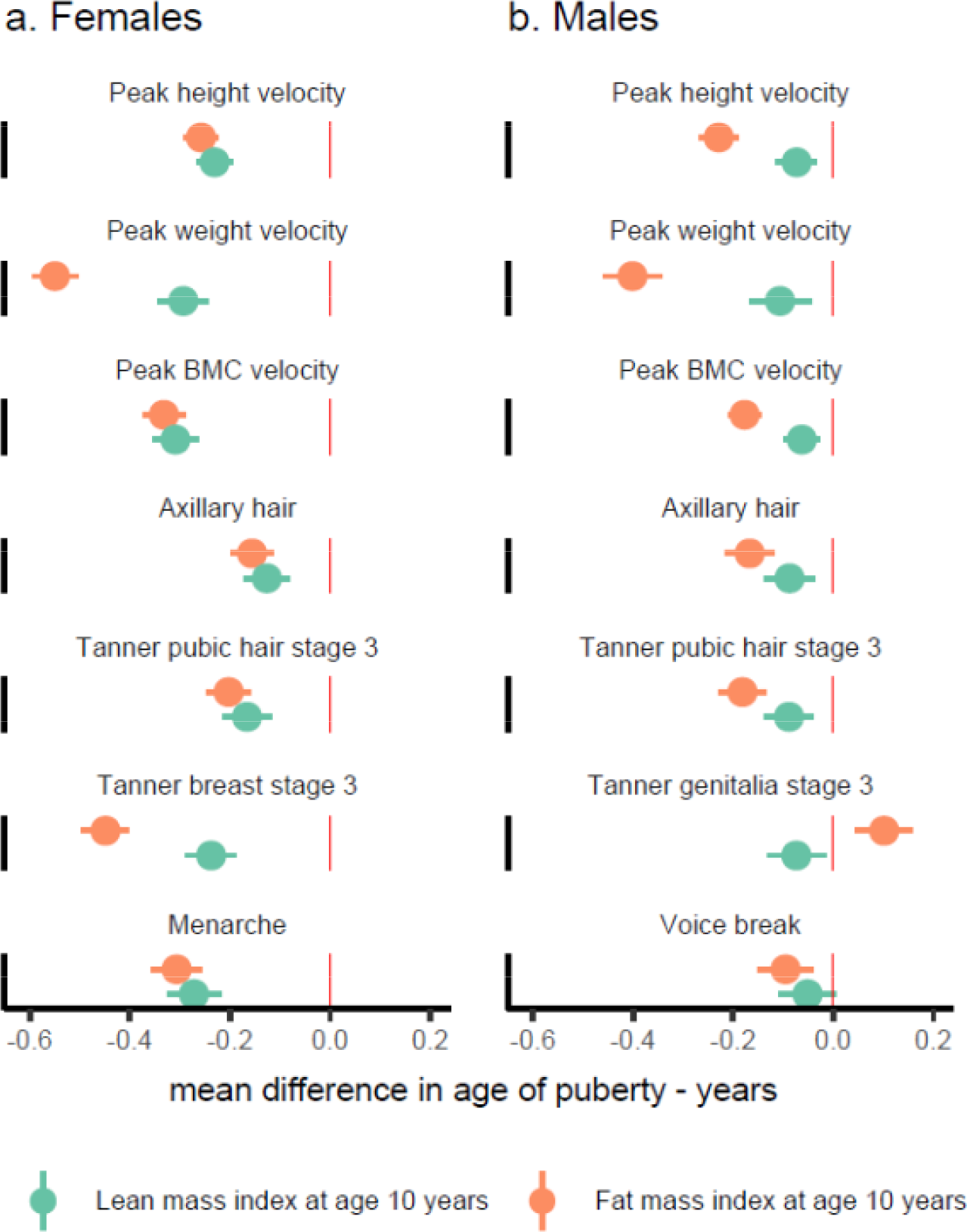
Association of pre-pubertal fat mass and lean mass with measures of pubertal age. Figure shows mean difference in age of each puberty measure per standard deviation higher pre-pubertal fat mass and lean mass indices (measured at age 10 years). Estimates were obtained from separate multivariable linear regression models for each pubertal age measure and fat mass or lean mass, with adjustment for age at fat/lean mass assessment, childhood dietary intake, and maternal age at birth, maternal body mass index, maternal education, maternal smoking, and parity. Horizontal bars represent 95% confidence intervals.

## DISCUSSION

We used repeated assessments from a population-based cohort to examine and compare nine growth and development-based measures of puberty timing. We found that breast, pubic hair, and genitalia development were relatively early indicators of pubertal stage, whilst peak bone accrual, menarche, and voice breaking were later indicators. All pubertal age measures were interrelated, as demonstrated by positive phenotypic and genetic correlations. GRSs based on large scale GWAS’s for age at menarche and voice breaking were positively associated with all other pubertal age measures, and GRS’s for adulthood and childhood BMI were inversely associated with pubertal age measures. Pre-pubertal fat mass and lean mass were inversely associated with all pubertal age measures, the only exception a positive association between fat mass and genitalia stage in males. Whilst replication of our results in independent cohorts is needed, if confirmed, our findings suggest that this suit of nine measures could facilitate important research in birth cohorts where repeated assessments are common, e.g., to assess whether associations with risk factors or outcomes are comparable across all pubertal age measures or if they are specific to certain growth/development measures (and sex).

### Relation to previous studies

To the best of our knowledge, ours is the first study to investigate this collection of puberty timing measures. However, our findings are consistent with previous studies that examined some of the measures. These include a study of 14,000 participants in the Danish National Birth Cohort with repeat assessments of the six developmental measures (but none of the growth measures), obtained from the same parent-/self-report questionnaire that we used, which found that breast, genitalia, and pubic hair stages were early indicators of pubertal stage, with menarche and voice breaking late indicators^27^. Our findings on chronological sequence of measures were also consistent with results from the Edinburgh Longitudinal Growth Study where height, menarche, and clinical examinations of developmental stages were taken every half-year until 20 years in 74 females and 103 males^24^. That study also reported positive phenotypic correlations between these measures (r: 0.62 to 0.82 in males and 0.80 to 0.92 in females)^24^. Consistent sequence and phenotypic correlations to our study were also reported in a study conducted at 10 schools in the Copenhagen area on 730 males with 2-4 repeated clinical examinations of height and pubertal stages (r: 0.4 to 0.62)^28^.

One previous study of 2,000 US participants examined age of peak BMC velocity in relation to age of peak height velocity (but not other measures) and, consistent with our study, found that peak velocity occurred earlier for height than BMC in both females and males and that ages of both measures were positively correlated^29^. We found that mean age of peak velocity for height was similar or slightly younger than for weight, which agrees with findings from a study on 105 twin pairs^30^. Our finding of inverse associations between pre-pubertal fat mass and puberty timing is consistent with previous observations^28, 31, 32^. Our study adds to those previous studies by comparing associations across nine measures of pubertal timing, showing that this association is substantially stronger with peak weight velocity than other measures, and that childhood lean mass is also inversely associated with puberty timing. We could not identify previous studies that have examined the shared genetic architecture between pubertal timing measures, as done in our study.

### Interpretation of findings

The age sequence of the different puberty measures is broadly consistent with the underlying molecular and hormonal changes driving the appearance of these changes^2, 3, 33^. Our evidence of phenotypic and genetic correlations between measures suggest they may all be capturing the same process and have a shared heritable contribution (from common genetic variation). Likewise, the positive genetic correlations found between males and females for the various measures point to similar genetic factors driving pubertal timing in each sex. GRSs derived from published genome-wide significant SNPs for age at menarche and age at voice breaking associated positively with all other measures of pubertal age, which, along with the evidence of phenotypic and genetic correlations between measures, suggests they could all be used as measures of pubertal age when assessing determinants and consequences of puberty timing. Our multivariable regression results suggests that both higher fat mass and muscle mass in childhood might contribute to earlier puberty.

### Study limitations

Data on developmental measures were collected by questionnaire using parent/self-reporting which might result in larger measurement errors compared with growth measures, and these differences in measurement error might have biased observed differences in pubertal ages^34^. Assessment of Tanner stages was supported by pictorial depictions and accompanying explanations of each Tanner stage which might have mitigated against this^35^. Furthermore, studies that have used clinical assessments (i.e., observation by trained clinicians or research staff) rather than self-report have reported similar results to ours^24, 28^. Axillary hair was collected as a dichotomous response, which could result in an imprecise estimate of pubertal age. Only five repeated measures of BMC were available for deriving the age of peak BMC velocity which may have led to imprecise estimation^36^. GWAS sample sizes were small in ALSPAC which can lead to unstable LDSR genetic correlation estimates. Whilst analyses of prepubertal body composition were adjusted for measured confounders, we cannot rule out bias from residual or unmeasured confounding. ALSPAC participants were White Europeans and results might not generalise to other ethnic groups. Other pubertal age measures such as age of first ejaculation, and skeletal bone age were not available, and could have provided further information on puberty timing.

## Conclusion

Findings from this prospective population-based cohort study of males and females supported all nine growth and development-based measures pubertal age measures as useful measures of age at puberty, by providing evidence that they are measuring the same biological process. Choice of measure(s) to use in studies with plans for data collection is influenced by various factors, including research questions, available resources together with competing demands for other types of data to be collected, participant burden, and acceptability of data collection methods. For instance, studies comparing puberty timing between males and females could focus on the measures available in both sexes, such as height, weight, and BMC, as well as pubic or axillary hair.

Obtaining accurate measures requires repeat analyses whether using growth or developmental data, which can be challenging due to limited funding and research resources. Cohort studies that collect repeated data prospectively are research resources, often available to the global research community rather than funded to address a limited set of research questions. Thus, repeated height or weight data collections, which are likely to be relevant to many areas of study might become the basis for assessing pubertal age. However, there would be scientific value in other studies measuring as many of the measures we present so our finding might be replicated in independent studies. Further, availability of multiple measures would allow the comparison risk factors and outcome across pubertal age measures. Finally, the correlations presented here may be useful for harmonising measures across studies (e.g., meta-analysis).

## METHODS

This study was conducted using data from the ALSPAC cohort. A pre-specified analysis plan for this study is available at https://osf.io/3qndg/.

### Cohort description

ALSPAC is a multigenerational prospective birth cohort study that recruited pregnant women residing within the catchment area of three National Health Service authorities in southwest England with an expected date of delivery between April 1991 and December 1992^20–22^. The initial number of pregnancies enrolled was 14,541. Of these initial pregnancies, there was a total of 14,676 fetuses, resulting in 14,062 live births and 13,988 children who were alive at 1 year of age. When children were approximately 7 years old, an attempt was made to bolster the initial sample with eligible new cases. Total sample size for analyses using data collected after age 7 years was 15,447 pregnancies, and 15,658 offspring. Of these 14,901 were alive at 1 year of age. Detailed data have been collected from offspring and parents by questionnaires, data extraction from medical records, data linkage to health records, and dedicated clinic assessments.

ALSPAC participants provided written informed consent or assent for all measurements. Ethical approval for the ALSPAC study was obtained from the ALSPAC Law and Ethics Committee and the Local Research Ethics Committees. Consent for biological samples has been collected in accordance with the Human Tissue Act (2004). Details of all available data can be found in the ALSPAC study website which includes a fully searchable data dictionary and variable search tool (http://www.bristol.ac.uk/alspac/researchers/our-data/).

### Measurements

#### Puberty data collection from research clinics and questionnaires

Anthropometric and developmental puberty data (and pubertal age measures derived from these) are described in **Supplementary Table 1**. All participants were invited to attend nine repeated research clinic examinations from ages 7 to 17 years where their height (in cm) and weight (in kg) were measured. In five of the clinics (from ages 9 to 17 years), all participants underwent whole-body DXA scans from which total-body (less head) bone mineral content (BMC; in grams) was extracted. Exact age in months at completing each research clinic assessment was recorded.

Questionnaires on pubertal development (the ‘Growing and Changing Questionnaire’) were mailed to all participants on nine occasions from ages 8 to 17 years (**Supplementary Table 1**). Questionnaires could be answered by either parent or guardian, child, or a combination; over 70% of the first five questionnaires were completed with help from a parent or guardian and the last four questionnaires were mostly completed by the child alone (**Supplementary Table 5**). Each questionnaire collected information relating to the five Tanner stages of pubic hair, breasts (girls), and genitalia (boys) development using line drawings representing each stage with accompanying description (**Supplementary Fig. 1**). Each questionnaire collected data on the onset of menstruation in girls, and all except the first questionnaire collected data on changes in voice (boys). The last seven questionnaires (from ages 10 to 17 years) gathered data on the development of axillary hair. Exact age in months at completing each puberty questionnaire was recorded.

#### Genotyping and imputation

Children were genotyped using the Illumina HumanHap550 quad chip genotyping platform (Illumina) by 23andMe subcontracting the Wellcome Trust Sanger Institute (Cambridge, UK) and the Laboratory Corporation of America (Burlington, NC, USA). Raw genome-wide data were subjected to standard quality control methods. Individuals were excluded based on sex mismatches, minimal or excessive heterozygosity, disproportionate missingness (>3%), and insufficient sample replication (identity by descent (IBD)lJ<lJ0.8). Individuals of non-European ancestry were removed because source GWAS (described below) for puberty measures were conducted primarily in European populations. Single nucleotide polymorphisms (SNPs) with minor allele frequency <lJ1%, call rate <95%, or with evidence for violations of Hardy-Weinberg equilibrium (*P*<5×10^−7^) were removed. Cryptic relatedness was measured as proportion of IBD >lJ0.1. Related individuals that passed quality control thresholds were retained in subsequent phasing and imputation.

In total, 9,115 children and 500,527 SNPs passed quality control filters. Of these, 477,482 SNP genotypes in common between the sample of ALSPAC children and mothers were combined for imputation to the Haplotype Reference Consortium (HRCr1.1, 2016) panel. SNPs with genotype missingness >1% (11,396 SNPs) were removed prior to imputation. A further 321 subjects were removed due to ID mismatches. HRC panel was phased using ShapeIt (v2.r644) which utilizes relatedness during phasing, and imputation was performed using the Michigan imputation server. This resulted in 8,237 children with genotype data after exclusion of related subjects using cryptic relatedness measures described previously.

#### GRSs for female and male puberty timing, and adulthood and childhood BMI

GRSs were created using genome-wide significant SNPs from European ancestry GWAS meta-analyses on reported age at menarche^8^ and age at voice breaking^9^, and measured BMI in adulthood^26^ and childhood (age range from 3 to 10 years)^25^. The scores were calculated using 351 SNPs associated with age at menarche, 73 SNPs associated with age at voice breaking, 95 SNPs associated with adulthood BMI (2/97 SNPs were not available in ALSPAC), and 15 SNPs associated with childhood BMI. Scores were constructed by multiplying the number of effect alleles (or probability of effect alleles if imputed) at each SNP (0, 1, or 2) by its weighting, summing them, and dividing by the total number of SNPs used, and thus reflect the average per-SNP effect on their respective trait (age at menarche, age at voice breaking, adulthood BMI, or childhood BMI). All scores were standardised (to mean=0 and SD=1) prior to analysis (**Supplemental Fig. 2**).

#### Pre-pubertal body composition measurements and confounders

Pre-pubertal fat mass index (total body fat mass divided by height^2^) and lean mass index (total body lean mass divided by height^2^), both in units of kg/m^2^, were derived from DXA scans at mean age 9.9 years (exact age in months when scan was recorded). Measures were standardised by age and sex (to mean=0 and SD=1) prior to examining their association with pubertal age measures. Maternal age at birth, parity, maternal early pregnancy BMI, maternal education, maternal pregnancy smoking, and childhood diet were hypothesised to confound these associations and selected for inclusion as model adjustments. The confounders were reported in questionnaires in pregnancy for maternal factors and in parent-completed food frequency questionnaires for the child’s dietary intake when they were around 7 years old (total energy intake in kilojoules per day).

### Statistical analyses

#### Derivation of pubertal age measures

Data used to derive pubertal ages were collected prospectively using nine repeated research clinic assessments and nine puberty-specific questionnaires (**Fig 1**, **Supplementary Table 1**, **Supplementary Table 2**). Estimated puberty ages were analysed in months for all measures and presented in years to aid interpretation. All analyses were restricted to White ethnicity individuals (>95% of all participants) to enable consistency across phenotypic and genetic analyses. Analyses were performed in R version 4.02 (R Project for Statistical Computing).

Age at menarche was calculated as the first reported age at onset of menstruation. Pubertal age from height was derived using SITAR (Super Imposition by Translation And Rotation) models^23^. SITAR is a shape invariant nonlinear mixed effects model that fits a mean natural spline growth curve and tailors it by shifting and scaling (with three random effects) to obtain each individual (subject-specific) curve. These three random effects describe the *size*, *timing*, and *intensity* of individual growth relative to the mean growth curve. *Size* reflects up or down shifts in the mean curve, *timing* reflects left to right shifts in the mean curve, and *intensity* reflects shrinking or stretching of the age scale which rotates the mean curve^23^. Because SITAR assumes constant or zero growth post-puberty, pubertal age from weight and BMC (whose growth continues into adulthood) was derived using SITAR models with a fourth ‘*post-growth’* random effect to allow post pubertal growth to vary between individuals.

SITAR models were fitted separately in males and females on participants with at least one height, weight, or BMC measurement. The best fitting models were identified by comparing models with 2 to 5 knots (at quantiles of the age distribution) in the mean spline curve and inspecting the mean curves and Bayesian information criterion (BIC) values for each model (**Supplementary Table 6, Supplementary Fig 3**). Covariances for the random effects were modelled (**Supplementary Table 7**). Age at puberty was estimated using the *timing* random effect, with the three derived variables representing ages at peak growth velocity for height, weight, and BMC.

Puberty age from the five Tanner stages for pubic hair, breast, and genitalia development, the three voice breaking groups, and the binary responses on absence or presence of axillary hair were derived using nonlinear mixed effects models based on SITAR with up to two random effects for *timing* and *intensity*^24^. This was because all individuals are measured on the same five-point scale (or three for voice breaking, and two for axillary hair), so their position on the scale at any particular time depends purely on their developmental age at that time, taking into account their *timing* and *intensity* random effects. Models were fitted separately in males and females on participants with at least one questionnaire response. Where a model failed to converge, it was refitted using one random effect (for *timing*). Covariances were modelled when more than one random effect was included (**Supplementary Table 7**).

A similar approach to that used for growth phenotypes was used to identify the best fitting models (**Supplementary Table 6, Supplementary Fig 4**) and estimate pubertal age from these. The five derived age variables represent age in Tanner stage 3 of pubic hair, breast (females only), and genitalia (males only) development, and ages at voice breaking and axillary hair appearance. Because regression modelling can allow for measurement error, inconsistent responses (i.e., reporting a developmental stage that was lower than that reported in a previous questionnaire) were included in the analysis, except for inconsistent responses in voice breaking which were removing prior to modelling due to convergence issues.

#### Sequence, interrelationships, and shared genetic architecture between puberty timing measures

The age and chronological sequence of the derived puberty age measures were summarised using mean and standard deviation (SD). The time taken to transition between the different puberty measures was summarised by subtracting mean age of each puberty measure from mean age of the preceding measure. Bivariate scatterplots and pairwise phenotypic Pearson correlations were used to examine interrelationships between pubertal age measures.

Shared genetic associations between puberty age measures both within and between sexes was assessed using LDSR to estimate genetic correlations between pubertal age measures, using full GWAS summary statistics^37^. Summary data were obtained from a published GWAS for age at menarche^8^ (n=252,000) and were used. Summary data for all other measures (including voice break because full summary data were not available from the GWAS on age at voice break) were generated in ALSPAC (coded in years). For ALSPAC, linear regression was used to run each GWAS using BOLT-LMM (without adjustment for principal components as all the participants were from a small geographically defined region, with 96% of parents reporting they were White British). A reference map from BOLT-LMM was used to interpolate genetic map coordinates from each SNP physical (base pair) position. Reference LD scores from BOLT-LMM appropriate for the analysis of European-ancestry samples were used to calibrate BOLT-LMM. LD scores were matched to SNPs by base pair coordinate. GWAS was performed separately for male and female measures, and results for shared pubertal age measures (i.e., height, weight, BMC, pubic hair, and axillary hair) were meta-analysed using GWAMA. LDSR genetic correlations can be, and were obtained for all age at puberty measures in both sexes (i.e., even the sex specific ones).

#### Associations of GRSs for pubertal age and BMI with pubertal age measures

To assess the usefulness of our nine derived puberty age measures in respect to their strength of association with genetic predisposition to puberty timing and BMI, separate univariable linear regression models were used to examine associations of female and male puberty timing GRSs and adulthood and childhood BMI GRSs with each puberty age measure.

#### Association of pre-pubertal body composition with pubertal age measures

Effect of pre-pubertal body composition in terms of DXA-derived fat mass and lean mass indices (at age 10 years) on pubertal age measures was examined in separate multivariable linear regression models adjusted for exact age at measurement of fat mass and lean mass, and confounders (maternal age at birth, maternal education, parity, maternal early pregnancy BMI, maternal pregnancy smoking, and childhood dietary intake). DXA measures recorded after age of puberty were removed, leaving up to 2,491 females and 2,500 males for analysis. Fat mass and lean mass were coded in age- and sex-specific SD units (mean=0 and SD=1).

## Data Availability

Data used in this study were from the ALSPAC birth cohort.Researchers interested in accessing ALSPAC data used in this study will need to submit a research proposal (https://proposals.epi.bristol.ac.uk/) for consideration by the ALSPAC Executive Committee (managed access).

http://www.bristol.ac.uk/alspac/

## ACKNOWLEDGEMENTS

We are extremely grateful to all the families who took part in this study, the midwives for their help in recruiting them and the whole ALSPAC team, which includes interviewers, computer and laboratory technicians, clerical workers, research scientists, volunteers, managers, receptionists, and nurses. ALSPAC data were collected and managed using REDCap (Research Electronic Data Capture) electronic data capture tools hosted at the University of Bristol. GWAS data was generated by Sample Logistics and Genotyping Facilities at Wellcome Sanger Institute and LabCorp (Laboratory Corporation of America) using support from 23andMe.This project has received funding from the European Union’s Horizon 2020 research and innovation programme under grant agreement No. 874739 (LongITools). AE and DAL receive part of their salary from the European Union’s Horizon 2020 research and innovation program under grant agreement No. 101021566 (ART-HEALTH). AE, MF, AGS, JAB, JH, LDH, KT, NJT, and DAL work in a Unit that receives funds from the University of Bristol and UK Medical Research Council (MC_UU_00032/05 and MC_UU_00032/02). DAL is a National Institute of Health Research Senior Investigator (NF- 0616-10102) and is also supported by a British Hear Foundation Chair (CH/F/20/90003). The UK Medical Research Council and Wellcome (Grant ref: 217065/Z/19/Z), and the University of Bristol provide core support for ALSPAC. A comprehensive list of grants funding is available on the ALSPAC website (http://www.bristol.ac.uk/alspac/external/documents/grant-acknowledgements.pdf). The funders had no role in the design and conduct of the study; collection, management, analysis, and interpretation of the data; preparation, review, or approval of the manuscript; and decision to submit the manuscript for publication. AE had full access to all the data in the study and takes responsibility for the integrity of the data and accuracy of the data analysis.

## AUTHOR CONTRIBUTIONS

AE developed the idea for the paper with initial input from DAL and further input from all authors. AE developed the analysis plan with input from all authors. AE did the majority of the statistical analysis, with support from all authors. MF did the LDSR genetic correlation analysis. AGS calculated the GRS for age at voice break. JAB calculated the GRS for age at menarche, adulthood BMI, and childhood BMI. TJC, JH, and KT provided advice on fitting mixed effects models to estimate age of puberty. AE wrote the first draft of the manuscript. All authors provided feedback on the draft and approved the final version for submission.

## MATERIALS & CORRESPONDENCE

Ahmed Elhakeem, MRC Integrative Epidemiology Unit at the University of Bristol, Oakfield House, Oakfield Grove, Bristol BS8 2BN, UK.

## DATA AVAILABILITY

Researchers interested in accessing ALSPAC data used in this study will need to submit a research proposal (https://proposals.epi.bristol.ac.uk/) for consideration by the ALSPAC Executive Committee (managed access).

## CODE AVAILABILITY

Statistical code (and analysis plan) used for this paper can be found in the Open Science Framework website at https://osf.io/3qndg/.

