## Supplemental Material for "Evaluation and comparison of nine growth- and development-based measures of pubertal timing"

### **Supplementary Table 1** Description of the repeated pubertal growth and development data and the puberty age measures derived from these.

| **Data type (number of repeated measures)** | **Description and source of the prospective repeated puberty data collections** | **Definition of the derived measures of pubertal age (months), method** |
| --- | --- | --- |
| Height (nine repeated measures) | Measured to the nearest 0.1 cm using a Harpenden stadiometer during nine research clinics from mean age 7.6 to 17.8 years by accredited fieldworkers | age at peak height velocity,  mixed effects model |
| Weight (nine repeated measures) | Measured to the nearest 0.1 kg using a Tanita Body Fat Analyser during nine research clinics from mean age 7.6 to 17.8 years by accredited fieldworkers | age at peak weight velocity,  mixed effects model |
| Bone mass (five repeated measures) | Measured by Lunar Prodigy Dual-energy X-ray Absorptiometry (DXA) scans as total body (less head) bone mineral content (BMC; grams) during five research clinics from mean age 9.9 to 17.8 years by accredited fieldworkers | age at peak BMC velocity,  mixed effects model |
| Tanner pubic hair development stage (nine repeated measures) | Reported by parent and/or child during nine puberty questionnaires from mean age 8.2 to 17.0 years, by using line drawings with accompanying descriptions of the five Tanner stages of pubic hair development to identify the stage most closely matched to the child’s current stage. | age in Tanner stage 3 of pubic hair development,  mixed effects model |
| Tanner breast development stage (nine repeated measures) | Reported by parent and/or child during nine puberty questionnaires from mean age 8.2 to 17.0 years, by using line drawings with accompanying descriptions of the ﬁve Tanner stages of breast development to identify the stage most closely matched to the child’s current stage. | age in Tanner stage 3 of breast development,  mixed effects model |
| Tanner genitalia development stage (nine repeated measures) | Reported by parent and/or child during nine puberty questionnaires from mean age 8.2 to 17.0 years, by using line drawings with accompanying descriptions of the ﬁve Tanner stages of genitalia development to identify the stage most closely matched to the child’s current stage. | age in Tanner stage 3 of genitalia development,  mixed effects model |
| Menstruation (nine repeated measures) | Reported by parent and/or child during nine puberty questionnaires from mean age 8.2 to 17.0 years, in response to questions on whether the daughter’s ﬁrst menstrual period occurred and if it has, the month and year it started. | age at menarche,  the first reported age at onset of menstruation |
| Voice change (eight repeated measures) | Reported by parent and/or child during eight puberty questionnaires from mean age 9.7 to 17.0 years, in response to a question on whether the son’s voice (1) has changed and if it has, whether it was (2) occasionally a lot lower or if the voice had (3) changed totally. | age at voice break,  mixed effects model |
| Axillary hair (seven repeated measures) | Reported by parent and/or child during seven puberty questionnaires from mean age 9.7 to 17.0 years, in response to a question on whether the child had (1) started growing hair in the armpits or (2) not yet started. | age at axillary hair,  mixed effects model |

### **Supplementary Table 2.** Number and age of study participants that completed each research clinic assessment and puberty questionnaire

|  | Females | |  | Males | |
| --- | --- | --- | --- | --- | --- |
|  | N | mean age (SD) |  | N | mean age (SD) |
| *Research Clinic (C): height, weight, and bone mineral content (BMC) assessments* |  |  |  |  |  |
| C1 (height, weight) | 4,047 | 7.6 (0.3) |  | 4,170 | 7.6 (0.3) |
| C2 (height, weight) | 3,576 | 8.7 (0.3) |  | 3,598 | 8.7 (0.3) |
| C3 (height, weight, BMC) | 3,865 | 9.9 (0.3) |  | 3,772 | 9.9 (0.3) |
| C4 (height, weight) | 3,785 | 10.7 (0.3) |  | 3,699 | 10.7 (0.3) |
| C5 (height, weight, BMC) | 3,611 | 11.8 (0.2) |  | 3,496 | 11.8 (0.2) |
| C6 (height, weight) | 3,455 | 12.8 (0.2) |  | 3,320 | 12.9 (0.2) |
| C7 (height, weight, BMC) | 3,118 | 13.9 (0.2) |  | 3,009 | 13.9 (0.2) |
| C8 (height, weight, BMC) | 2,856 | 15.5 (0.4) |  | 2,572 | 15.5 (0.3) |
| C9 (height, weight, BMC) | 2,851 | 17.8 (0.5) |  | 2,217 | 17.8 (0.4) |
| N with ≥1 height assessment | 4,186 | - |  | 4,251 | - |
| N with ≥1 weight assessment | 4,183 | - |  | 4,248 | - |
| N with ≥1 BMC assessment | 3,747 | - |  | 3,680 | - |
| *Puberty Questionnaire (Q): Tanner pubic hair, breast, and genitalia stages, and axillary hair, voice break, and menarche data collection* |  |  |  |  |  |
| Q1 (no axillary hair or voice break) | 3,298 | 8.2 (0.3) |  | 2,947 | 8.2 (0.3) |
| Q2 (no axillary hair) | 3,652 | 9.7 (0.1) |  | 3,357 | 9.7 (0.1) |
| Q3 (no axillary hair) | 3,482 | 10.7 (0.1) |  | 3,156 | 10.7 (0.1) |
| Q4 (all) | 3,331 | 11.7 (0.1) |  | 2,991 | 11.7 (0.1) |
| Q5 (all) | 3,195 | 13.1 (0.2) |  | 2,871 | 13.1 (0.2) |
| Q6 (all) | 2,868 | 14.7 (0.1) |  | 2,286 | 14.7 (0.1) |
| Q7 (all) | 2,565 | 15.4 (0.3) |  | 2,292 | 15.4 (0.3) |
| Q8 (all) | 2,813 | 16.1 (0.1) |  | 1,938 | 16.1 (0.1) |
| Q9 (all) | 2,600 | 17.0 (0.1) |  | 1,762 | 17.0 (0.1) |
| N with ≥1 Tanner pubic hair response | 4,276 | - |  | 4,074 | - |
| N with ≥1 Tanner genitalia response | - | - |  | 4,041 | - |
| N with ≥1 Tanner breasts response | 4,273 | - |  | - | - |
| N with ≥1 axillary hair response | 4,031 | - |  | 3,804 | - |
| N with ≥1 voice break response | - | - |  | 4,020 | - |
| N with data on age at menarche | 3,457 | - |  | - | - |

### **Supplementary Table 3.** Comparison of the included study participants with data from at least one puberty questionnaire or research clinic with those excluded due to missing data.

|  | Males | | Females | |
| --- | --- | --- | --- | --- |
|  | Included (4,251) | Excluded (1,260) | Included (4,276) | Excluded (975) |
| Maternal pregnancy smoking [N (%)] |  |  |  |  |
| No | 3729 (95.8) | 408 (90.7) | 3750 (94.9) | 278 (93.3) |
| Yes | 164 (4.2) | 42 (9.3) | 200 (5.1) | 20 (6.7) |
| Maternal education [N (%)] |  |  |  |  |
| CSE | 588 (14.4) | 158 (29.8) | 596 (14.6) | 106 (30.1) |
| Vocational | 380 (9.3) | 73 (13.8) | 378 (9.3) | 37 (10.5) |
| O level | 1467 (35.8) | 183 (34.5) | 1429 (35.0) | 126 (35.8) |
| A level | 1054 (25.7) | 81 (15.3) | 1037 (26.4) | 57 (16.2) |
| Degree | 608 (14.8) | 35 (6.6) | 641 (15.7) | 26 (7.4) |
| Maternal parity [N (%)] |  |  |  |  |
| 0 | 1808 (45.2) | 201 (39.7) | 1820 (45.7) | 138 (41.1) |
| 1 | 1425 (35.6) | 187 (37.0) | 1469 (36.9) | 111 (33.0) |
| 2 | 572 (14.3) | 77 (15.2) | 526 (13.2) | 60 (17.9) |
| 3 or more | 198 (5.0) | 41 (8.1) | 170 (4.3) | 27 (8.0) |
| Maternal pregnancy BMI – kg/m^2^ [mean (SD)] | 23.0 (3.8) | 22.9 (3.8) | 22.9 (3.8) | 23.0 (4.2) |
| Maternal age at birth – years [mean (SD)] | 29.1 (4.6) | 27.0 (4.9) | 28.9 (4.5) | 27.1 (4.7) |
| Child energy intake – kJ/day [mean (SD)] | 7780 (1832) | 7901 (2011) | 7516 (1727) | 8277 (2068) |

### **Supplementary Table 4.** Genetic correlation estimates (using LDSR) between pairs of puberty age measures.

| **Puberty age measure 1 (n)** | **Puberty age measure 2 (n)** | **Estimate** | **SE** | ***P*** |
| --- | --- | --- | --- | --- |
| Menarche (n=252,000) | Axillary hair - females (n=3,161) | 0.339 | 0.084 | 5 x 10^-05^ |
| Menarche (n=252,000) | Axillary hair - females and males (n=6,175) | 0.713 | 0.263 | 0.007 |
| Menarche (n=252,000) | Peak BMC velocity - females and males (n=6,228) | 1.189 | 0.389 | 0.002 |
| Menarche (n=252,000) | Peak BMC velocity - males (n=3,109) | 0.782 | 0.292 | 0.007 |
| Menarche (n=252,000) | Peak height velocity - females (n=3,357) | 1.014 | 0.223 | 5 x 10^-06^ |
| Menarche (n=252,000) | Peak height velocity - females and males (n=6,782) | 0.875 | 0.101 | 5 x 10^-18^ |
| Menarche (n=252,000) | Peak height velocity - males (n=3,425) | 0.526 | 0.104 | 5 x 10^-07^ |
| Menarche (n=252,000) | Tanner breast stage 3 (n=3,296) | 0.723 | 0.130 | 2 x 10^-08^ |
| Menarche (n=252,000) | Tanner pubic hair stage 3 - females (n=3,295) | 0.808 | 0.284 | 0.0045 |
| Menarche (n=252,000) | Tanner pubic hair stage 3 - females and males (n=6,471) | 0.639 | 0.094 | 1 x 10^-11^ |
| Menarche (n=252,000) | Tanner pubic hair stage 3 - males (n=3,176) | 0.541 | 0.159 | 0.0007 |
| Peak height velocity - females and males (n=6,782) | Axillary hair - females (n=3,161) | 0.384 | 0.202 | 0.06 |
| Peak height velocity - females and males (n=6,782) | Axillary hair - females and males (n=6,175) | 0.791 | 0.255 | 0.002 |
| Peak height velocity - females and males (n=6,782) | Peak BMC velocity - females and males (n=6,228) | 0.984 | 0.150 | 5 x 10^-11^ |
| Peak height velocity - females and males (n=6,782) | Peak BMC velocity - males (n=3,109) | 0.978 | 0.267 | 0.0003 |
| Peak height velocity - females and males (n=6,782) | Peak weight velocity - females (n=3,350) | 0.508 | 0.503 | 0.3 |
| Peak height velocity - females and males (n=6,782) | Tanner breast stage 3 (n=3,296) | 0.887 | 0.151 | 4 x 10^-09^ |
| Peak height velocity - females and males (n=6,782) | Tanner pubic hair stage 3 - females (n=3,295) | 0.990 | 0.288 | 0.0006 |
| Peak height velocity - females and males (n=6,782) | Tanner pubic hair stage 3 - females and males (n=6,471) | 0.959 | 0.136 | 1 x 10^-12^ |
| Peak height velocity - females and males (n=6,782) | Tanner pubic hair stage 3 - males (n=3,176) | 1.080 | 0.310 | 0.0005 |
| Peak height velocity - females and males (n=6,782) | Tanner genitalia stage 3 (n=,3,680) | 0.762 | 0.997 | 0.4 |
| Peak BMC velocity - females and males (n=6,228) | Axillary hair - females (n=3,161) | 0.173 | 0.331 | 0.6 |
| Peak BMC velocity - females and males (n=6,228) | Axillary hair - females and males (n=6,175) | 0.736 | 0.409 | 0.07 |
| Peak BMC velocity - females and males (n=6,228) | Peak height velocity - females (n=3,357) | 0.775 | 0.291 | 0.008 |
| Peak BMC velocity - females and males (n=6,228) | Peak height velocity - males (n=3,425) | 1.022 | 0.201 | 4 x 10^-07^ |
| Peak BMC velocity - females and males (n=6,228) | Peak weight velocity - females (n=3,350) | 0.210 | 0.847 | 0.8 |
| Peak BMC velocity - females and males (n=6,228) | Tanner breast stage 3 (n=3,296) | 0.699 | 0.299 | 0.02 |
| Peak BMC velocity - females and males (n=6,228) | Tanner pubic hair stage 3 - females (n=3,295) | 1.098 | 0.503 | 0.03 |
| Peak BMC velocity - females and males (n=6,228) | Tanner pubic hair stage 3 - males (n=3,176) | 1.397 | 0.486 | 0.004 |
| Tanner pubic hair stage 3 - females and males (n=6,471) | Axillary hair - females (n=3,161) | 0.465 | 0.204 | 0.02 |
| Tanner pubic hair stage 3 - females and males (n=6,471) | Axillary hair - females and males (n=6,175) | 0.680 | 0.237 | 0.004 |
| Tanner pubic hair stage 3 - females and males (n=6,471) | Peak BMC velocity - females and males (n=6,228) | 1.133 | 0.308 | 0.0002 |
| Tanner pubic hair stage 3 - females and males (n=6,471) | Peak BMC velocity - males (n=3,109) | 1.031 | 0.408 | 0.01 |
| Tanner pubic hair stage 3 - females and males (n=6,471) | Peak height velocity - females (n=3,357) | 0.717 | 0.186 | 0.0001 |
| Tanner pubic hair stage 3 - females and males (n=6,471) | Peak height velocity - males (n=3,425) | 0.985 | 0.197 | 6 x 10^-07^ |
| Tanner pubic hair stage 3 - females and males (n=6,471) | Peak weight velocity - females (n=3,350) | 1.062 | 1.201 | 0.4 |
| Tanner pubic hair stage 3 - females and males (n=6,471) | Tanner breast stage 3 (n=3,296) | 0.389 | 0.208 | 0.06 |
| Tanner pubic hair stage 3 - females and males (n=6,471) | Voice breaking (n=4,020) | 0.750 | 0.972 | 0.4 |
| Axillary hair - females and males (n=6,175) | Peak BMC velocity - males (n=3,109) | 0.627 | 0.487 | 0.2 |
| Axillary hair - females and males (n=6,175) | Peak height velocity - females (n=3,357) | 0.827 | 0.382 | 0.03 |
| Axillary hair - females and males (n=6,175) | Peak height velocity - males (n=3,425) | 0.714 | 0.274 | 0.009 |
| Axillary hair - females and males (n=6,175) | Peak weight velocity - females (n=3,350) | 0.449 | 0.922 | 0.6 |
| Axillary hair - females and males (n=6,175) | Tanner breast stage 3 (n=3,296) | 0.820 | 0.375 | 0.03 |
| Axillary hair - females and males (n=6,175) | Tanner pubic hair stage 3 - males (n=3,176) | 0.281 | 0.406 | 0.5 |
| Peak height velocity - females (n=3,357) | Peak height velocity - males (n=3,425) | 0.635 | 0.317 | 0.05 |
| Peak height velocity - females (n=3,357) | Peak weight velocity - females (n=3,350) | 0.433 | 0.520 | 0.4 |
| Peak height velocity - females (n=3,357) | Tanner pubic hair stage 3 - males (n=3,176) | 0.800 | 0.431 | 0.06 |
| Peak height velocity - females (n=3,357) | Tanner genitalia stage 3 (n=,3,680) | 0.590 | 1.103 | 0.6 |
| Peak height velocity - males (n=3,425) | Peak weight velocity - females (n=3,350) | 0.598 | 0.878 | 0.5 |
| Peak height velocity - males (n=3,425) | Tanner pubic hair stage 3 - males (n=3,176) | 1.142 | 0.272 | 3 x 10^-05^ |
| Peak height velocity - males (n=3,425) | Tanner genitalia stage 3 (n=,3,680) | 0.781 | 0.941 | 0.4 |
| Tanner breast stage 3 (n=3,296) | Peak BMC velocity - males (n=3,109) | 0.525 | 0.408 | 0.2 |
| Tanner breast stage 3 (n=3,296) | Peak height velocity - females (n=3,357) | 0.999 | 0.154 | 8 x 10^-11^ |
| Tanner breast stage 3 (n=3,296) | Peak height velocity - males (n=3,425) | 0.657 | 0.274 | 0.02 |
| Tanner breast stage 3 (n=3,296) | Peak weight velocity - females (n=3,350) | 1.154 | 0.988 | 0.2 |
| Tanner breast stage 3 (n=3,296) | Tanner pubic hair stage 3 - females (n=3,295) | 0.509 | 0.279 | 0.07 |
| Tanner breast stage 3 (n=3,296) | Tanner pubic hair stage 3 - males (n=3,176) | 0.388 | 0.333 | 0.2 |
| Tanner pubic hair stage 3 - females (n=3,295) | Peak height velocity - females (n=3,357) | 0.731 | 0.265 | 0.006 |
| Tanner pubic hair stage 3 - females (n=3,295) | Peak height velocity - males (n=3,425) | 1.011 | 0.475 | 0.03 |
| Tanner pubic hair stage 3 - females (n=3,295) | Peak weight velocity - females (n=3,350) | 0.827 | 0.893 | 0.4 |
| Tanner pubic hair stage 3 - females (n=3,295) | Tanner pubic hair stage 3 - males (n=3,176) | 1.330 | 0.692 | 0.05 |
| Axillary hair - females (n=3,161) | Peak BMC velocity - males (n=3,109) | 0.179 | 0.378 | 0.6 |
| Axillary hair - females (n=3,161) | Peak height velocity - females (n=3,357) | 0.468 | 0.244 | 0.05 |
| Axillary hair - females (n=3,161) | Peak height velocity - males (n=3,425) | 0.301 | 0.226 | 0.2 |
| Axillary hair - females (n=3,161) | Peak weight velocity - females (n=3,350) | 0.261 | 0.680 | 0.7 |
| Axillary hair - females (n=3,161) | Tanner breast stage 3 (n=3,296) | 0.445 | 0.244 | 0.07 |
| Axillary hair - females (n=3,161) | Tanner pubic hair stage 3 - females (n=3,295) | 0.867 | 0.280 | 0.002 |
| Axillary hair - females (n=3,161) | Tanner pubic hair stage 3 - males (n=3,176) | 0.264 | 0.322 | 0.4 |
| Peak BMC velocity - males (n=3,109) | Peak height velocity - females (n=3,357) | 0.675 | 0.528 | 0.2 |
| Peak BMC velocity - males (n=3,109) | Peak height velocity - males (n=3,425) | 1.052 | 0.179 | 4 x 10^-09^ |
| Peak BMC velocity - males (n=3,109) | Peak weight velocity - females (n=3,350) | 0.592 | 1.184 | 0.6 |
| Peak BMC velocity - males (n=3,109) | Tanner pubic hair stage 3 - females (n=3,295) | 1.078 | 0.757 | 0.2 |
| Peak BMC velocity - males (n=3,109) | Tanner pubic hair stage 3 - males (n=3,176) | 1.200 | 0.438 | 0.006 |
| Voice breaking (n=4,020) | Tanner pubic hair stage 3 - females (n=3,295) | 0.789 | 1.567 | 0.6 |
| Voice breaking (n=4,020) | Tanner pubic hair stage 3 - males (n=3,176) | 0.705 | 0.948 | 0.5 |
| Voice breaking (n=4,020) | Peak weight velocity - females (n=3,350) | 0.635 | 1.586 | 0.7 |
| Voice breaking (n=4,020) | Tanner genitalia stage 3 (n=,3,680) | 0.604 | 1.482 | 0.7 |
| Tanner genitalia stage 3 (n=,3,680) | Tanner pubic hair stage 3 - males (n=3,176) | 0.425 | 0.657 | 0.5 |

Genetic correlations were estimated using LD score regression with summary GWAS data obtained from Day et al. for age at menarche and in ALSPAC for all other measures of pubertal age. Note, genetic correlation estimates are unbounded, so some results fall outside 1. If the true genetic correlation is near 1, and the estimated genetic correlation is the true value plus or minus some estimation error, it’s possible that the estimate (true genetic correlation + error) will be greater than 1. For many traits there was low sample size which can lead to unstable results (i.e., more likely to estimate genetic correlation > 1). When both the true genetic correlation and sample overlap are high, LD score regression can give exaggerated estimates (>1).

### **Supplementary Table 5.** Numbers of study participants that had help from parent/guardian with completing each puberty questionnaire

|  | Females | |  | Males | |
| --- | --- | --- | --- | --- | --- |
|  | N | % |  | N | % |
| Child had help completing puberty questionnaire (Q) |  |  |  |  |  |
| Q1 |  |  |  |  |  |
| Yes | 3241 | 99.0 |  | 2871 | 98.4 |
| No | 33 | 1.0 |  | 48 | 1.6 |
| Q2 |  |  |  |  |  |
| Yes | 3544 | 97.7 |  | 3107 | 96.5 |
| No | 82 | 2.3 |  | 114 | 3.5 |
| Q3 |  |  |  |  |  |
| Yes | 3379 | 97.8 |  | 2874 | 94.9 |
| No | 75 | 2.2 |  | 153 | 5.1 |
| Q4 |  |  |  |  |  |
| Yes | 2903 | 87.8 |  | 2311 | 80.9 |
| No | 404 | 12.2 |  | 545 | 19.1 |
| Q5 |  |  |  |  |  |
| Yes | 2500 | 79.2 |  | 1975 | 71.8 |
| No | 656 | 20.8 |  | 774 | 28.2 |
| Q6 |  |  |  |  |  |
| Yes | 404 | 14.3 |  | 246 | 11.0 |
| No | 2419 | 85.7 |  | 1988 | 89.0 |
| Q7 |  |  |  |  |  |
| Yes | 319 | 12.7 |  | 142 | 6.4 |
| No | 2199 | 87.3 |  | 2071 | 93.6 |
| Q8 |  |  |  |  |  |
| Yes | 250 | 9.0 |  | 141 | 7.4 |
| No | 2532 | 91.0 |  | 1769 | 92.6 |
| Q9 |  |  |  |  |  |
| Yes | 146 | 5.7 |  | 93 | 5.4 |
| No | 2427 | 94.3 |  | 1644 | 94.6 |

### **Supplementary Table 6.** BIC values from mixed effects models with different degrees of freedom (df)

|  | BIC | | | |
| --- | --- | --- | --- | --- |
|  | df=3  (2 knots) | df=4  (3 knots) | df=5  (4 knots) | df=6  (5 knots) |
| *Females* |  |  |  |  |
| Height | 114776 | 114369 | 114218 | **114132** |
| Weight | 143063 | 143068 | **142163** | 142746 |
| BMC | **165547** | 167436 | nc | 170841 |
| Tanner pubic hair stage | 38159 | **37177** | 37531 | 36911 |
| Tanner breast stage | **40363** | nc | 40783 | 40640 |
| Axillary hair | -10348 | -12226 | -20485 | **-27678** |
| *Males* |  |  |  |  |
| Height | nc | 116185 | 115770 | **115730** |
| Weight | 138753 | nc | **134206** | nc |
| BMC | 151301 | nc | **152763** | 154855 |
| Tanner pubic hair stage | 29076 | 29182 | 28998 | 29379 |
| Tanner genitalia stage | **44390** | 44812 | 45152 | 46142 |
| Axillary hair | -530 | -4982 | **-9733** | -8740 |
| Voice break | 15258 | 14995 | 14683 | **14619** |

nc=model did not converge. Selected models are highlighted in bold font.

### **Supplementary Table 7.** Correlations between the size, timing, intensity, and post-growth random effects from mixed effects models that included at least two of these random effects

|  | Correlations between random effects | | | | | |
| --- | --- | --- | --- | --- | --- | --- |
|  | timing and intensity | timing and size | size and intensity | timing and post-growth | size and post-growth | intensity and post-growth |
| *Females* |  |  |  |  |  |  |
| Height | 0.07 | 0.30 | 0.43 | - | - | - |
| Weight | 0.26 | 0.18 | 0.42 | -0.04 | 0.78 | -0.16 |
| BMC | 0.37 | 0.19 | 0.68 | -0.63 | 0.53 | -0.63 |
| Tanner pubic hair stage | -0.50 | - | - | - | - | - |
| Tanner breast stage | 0.19 | - | - | - | - | - |
| *Males* |  |  |  |  |  |  |
| Height | 0.15 | 0.37 | 0.56 |  |  |  |
| Weight | 0.75 | 0.49 | 0.55 | -0.11 | 0.68 | -0.15 |
| BMC | 0.21 | 0.37 | 0.56 | 0.35 | 0.93 | 0.23 |
| Tanner pubic hair stage | 0.66 | - | - | - | - | - |

Age modelled as log(age) for all measures except BMC in females

### **Supplementary Fig. 1** Instructions provided to the ALSPAC study participants for reporting Tanner stages using repeated puberty questionnaires from ages 7-17 years.

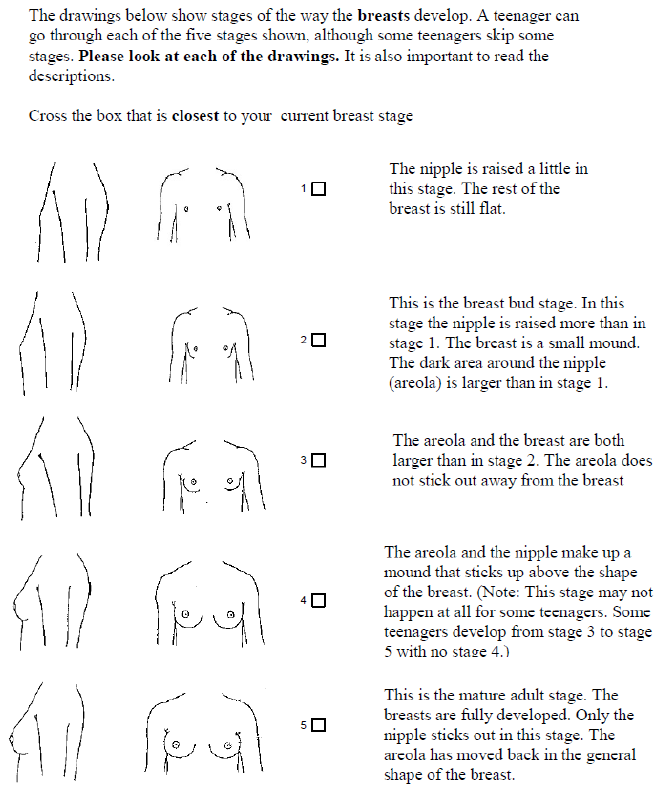

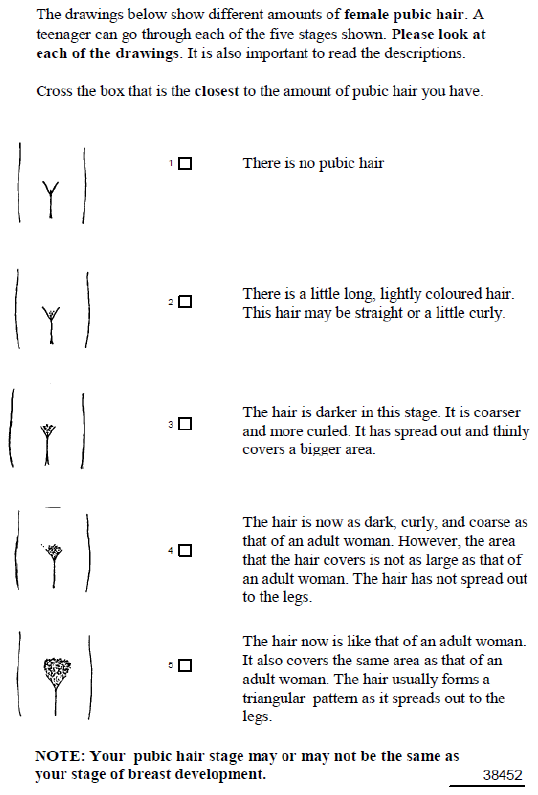

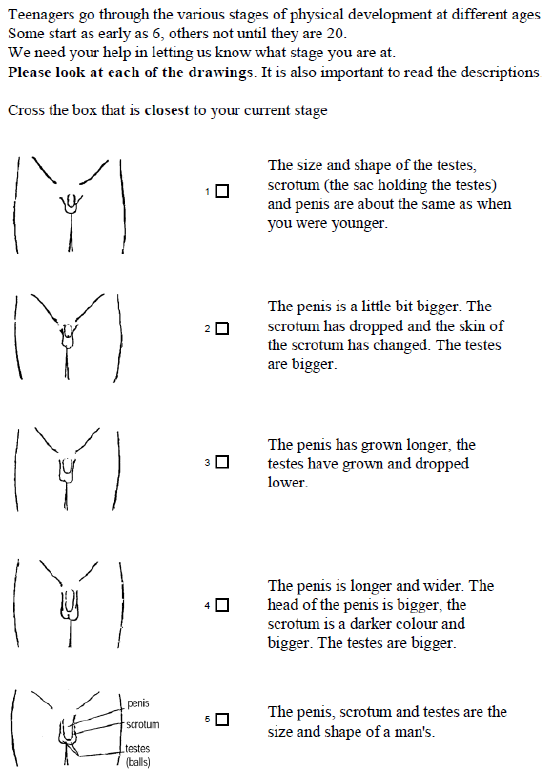

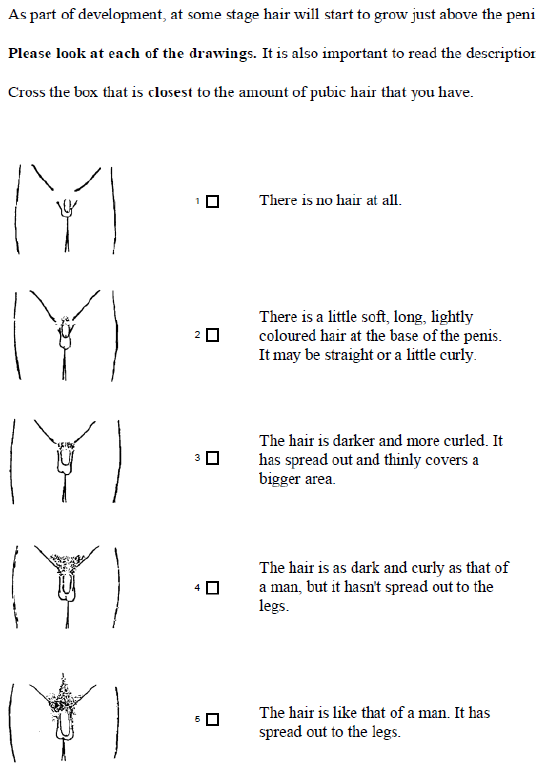

Figures reproduced with consent from the ALSPAC Executive

### **Supplementary Fig. 2** Distribution of standardized genetic risk scores

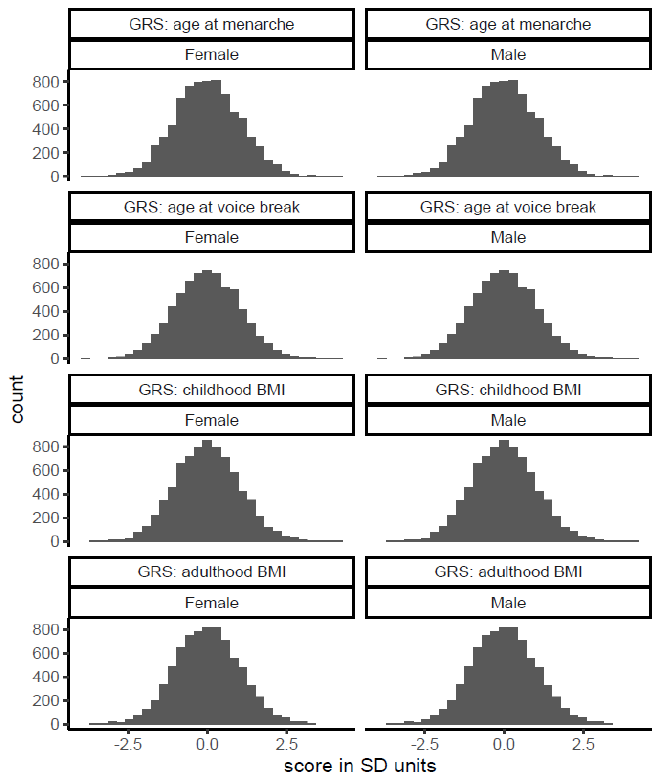

### **Supplementary Fig. 3** Estimated mean distance and velocity curves for height, weight, and BMC from the selected mixed effects models

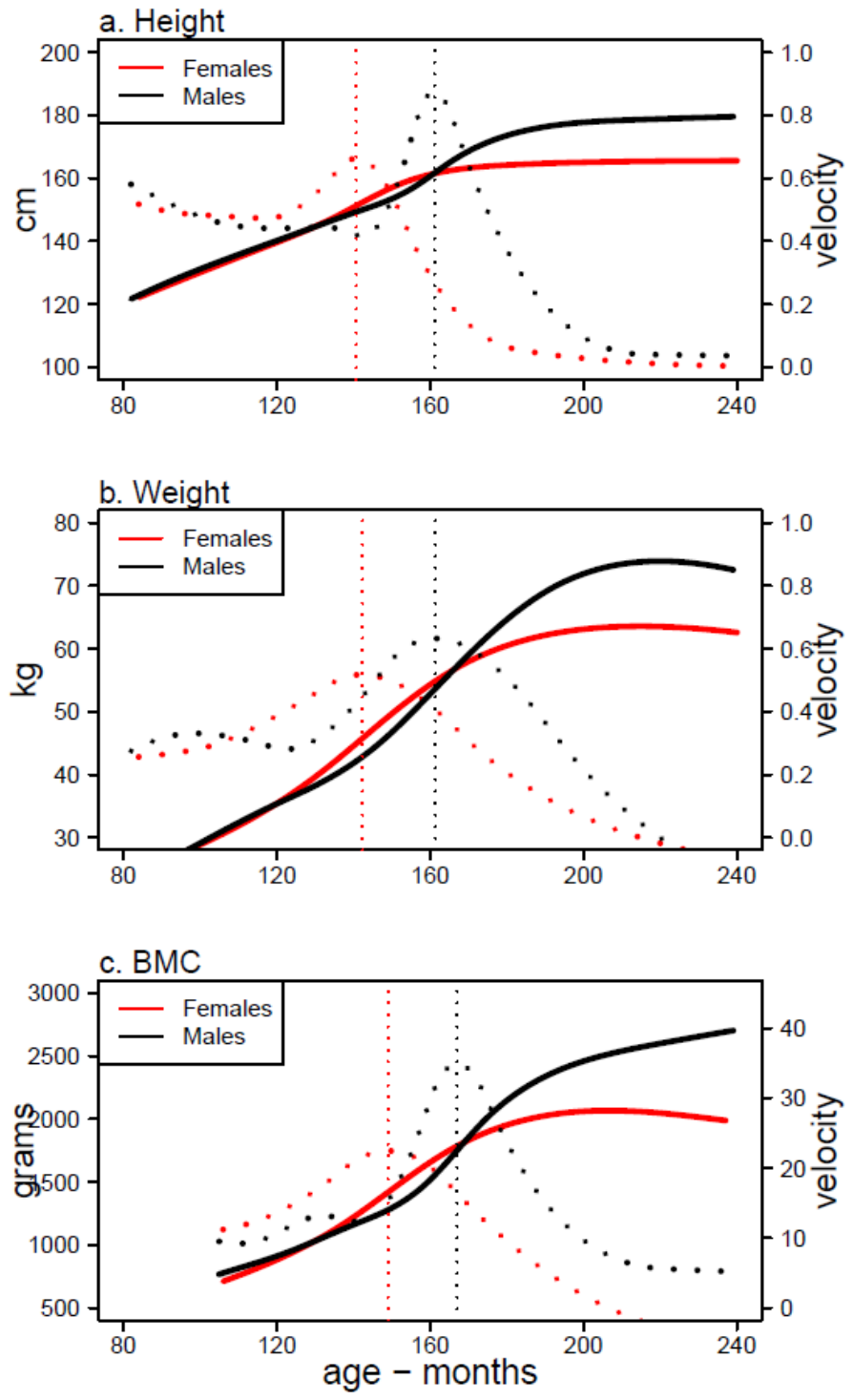

### **Supplementary Fig. 4** Estimated mean distance and velocity curves for Tanner stages, axillary hair, and voice break from the selected mixed effects models

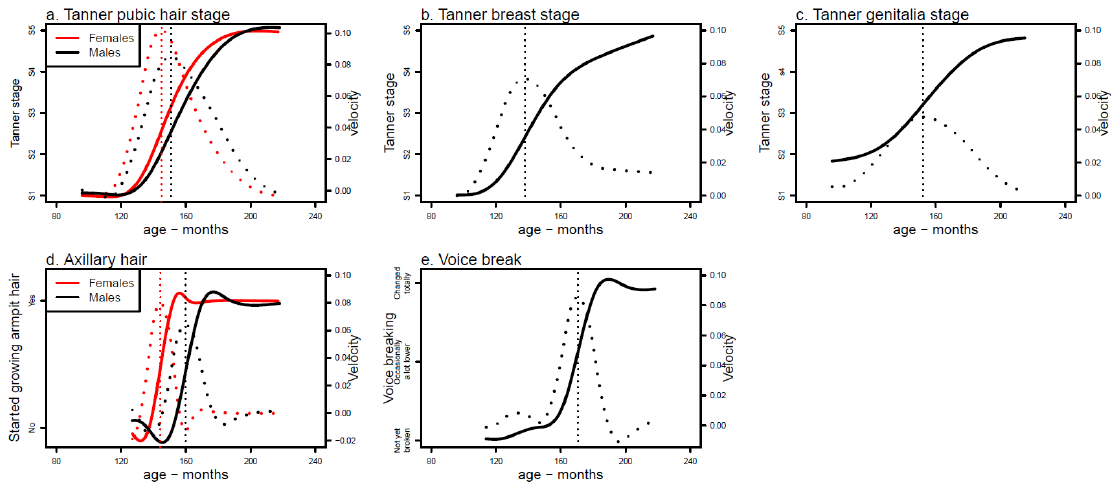
